# THE PROBOSCIS SENSORY INDEX PREDICTS VECTOR ANTHROPOPHILY AND HOST SELECTION IN NORTHERN NIGERIA

**DOI:** 10.64898/2026.09.07.26362427

**Authors:** Raphael Kawep Njapdze, Idorenyin Bassey Ekerette

## Abstract

**Background:** Unexplained spatial heterogeneity in field-observed Human Blood Index (HBI) hinders vector-borne disease control. We synthesized micro-morphological sensory structure densities across primary disease vectors to formulate a novel Proboscis Sensory Index (PSI) from micro-morphological sensory structure densities to evaluate its capacity to predict vector anthropophily, zooprophylactic modulation, and spatial transmission risk across Northern Nigeria.

**Methods:** A systematic review of 42 studies (N = 18,647 blood-fed mosquitoes across 126 georeferenced sites, 2005–2025) synthesized baseline species-specific PSI metrics from electron microscopy sensilla counts. A multi-variable binomial Generalized Linear Mixed Model (GLMM) evaluated interactions between PSI, livestock, human density, and environmental covariates. Predictive accuracy was validated against field-derived Entomological Inoculation Rates (EIR) and historical outbreak records.

**Results:** The overall pooled HBI was 0.642 (95% CI: 0.588 – 0.694), exhibiting significant variation across the Sudan (0.728), Guinea (0.615), and Sahel (0.584) savannas. Species-specific PSI values graded from Anopheles gambiae s.s. (8.91 ± 0.34), Anopheles funestus (8.12 ± 0.38), and Anopheles arabiensis (7.68 ± 0.41) to Culex quinquefasciatus (4.87 ± 0.52). In GLMM analysis, PSI was the strongest predictor of anthropophily (β = 0.89, aOR = 2.44, 95% CI: 2.32 – 2.56, p < 0.001), with fixed effects explaining 78% of total field variance (Marginal R^2^ = 0.78). High-PSI species maintained elevated anthropophily regardless of livestock availability, whereas low-PSI species displayed steep zooprophylactic diversion (ΔHBI = −0.71, p < 0.001) under high cattle density. Incorporating PSI reduced localized EIR prediction error by 64.2% (RMSE = 15.0 vs. 41.8 infective bites/person/month), and high-resolution (1 km2) predictive mapping correctly identified 87.1% (27/31) of historically documented outbreak Local Government Areas.

**Conclusion:** Micro-sensory organ density dictates vector host preference and livestock-mediated zooprophylactic potential. Integrating PSI into spatial models resolves localized transmission heterogeneity, guiding targeted vector control prioritizing indoor residual spraying and dual-ingredient nets in high-PSI foci, while leveraging endectocides and larval management in low-PSI pastoral corridors.

## INTRODUCTION

Vector-borne diseases, particularly malaria and emerging arboviruses (dengue, Zika, yellow fever), constitute the greatest infectious disease burden in sub-Saharan Africa. Nigeria alone accounts for 27 % of global malaria cases and 31 % of malaria deaths, with transmission intensity highest in the northern Sahelian and Sudan savanna zones (1,2). Despite scaled interventions, fine-scale spatial heterogeneity in transmission persists, driving recurrent localized outbreaks that evade blanket control strategies (3).

Predictive risk modeling has become essential for precision public health, guiding targeted deployment of indoor residual spraying, long-lasting insecticidal nets, and reactive vaccination. Current models in West Africa rely predominantly on classical entomological indices entomological inoculation rate (EIR), human biting rate (HBR), sporozoite rate, parity rate, and vector density often integrated with environmental covariates via geostatistical or machine-learning frameworks (4,5,6). However, these approaches assume uniform infectiousness of mosquito bites and fail to incorporate extreme short-range heterogeneity in host preference observed in major vectors such as *Anopheles gambiae* s.l. and *Aedes aegypti* across northern Nigeria (7,8,9,10,11).

Multiple studies have documented dramatic shifts from strongly anthropophilic to zoophilic feeding within 20–100 km, modulated by cattle density, settlement patterns, and nocturnal human outdoor activity. This proboscis-mediated host selection detected through forensic blood-meal analysis, human blood index (HBI), and host choice indices is almost never integrated into predictive models (12,13,14,15). The mosquito proboscis itself plays a pivotal sensory role: thermosensory neurons in the distal labellum detect radiant heat and skin temperature gradients via TRPA1 channels, while olfactory sensilla simultaneously perceive CO□, lactic acid, ammonia, and other host volatiles, synergising at close range to refine host discrimination and probing decisions (16,17). Ignoring this sensory-driven feeding plasticity leads current models to over- or underestimate localized transmission intensity and outbreak risk by factors of 3–10×, particularly in cattle-rich Sahelian communities where zooprophylaxis or zoopotentiation effects dominate (18,19,20).

This study therefore aimed to develop and validate a novel spatially explicit predictive modeling framework that incorporates proboscis-mediated host preference heterogeneity (via forensic blood-meal analysis and HBI gradients) alongside classical entomological indices and livestock density covariates at sub-LGA resolution in northern Nigeria.

Specific objectives were:

1. to quantify fine-scale spatial variation in anthropophilic/zoophilic feeding behaviour across Sahelian and Sudan savanna zones using existing and new forensic blood-meal data;
2. to integrate host preference gradients with entomological inoculation rates to derive corrected vectorial capacity and basic reproduction number (R□) estimates;
3. to generate high-resolution predictive risk maps that explicitly account for cattle-mediated modulation of vector–human contact; and
4. to evaluate model performance against standard entomological-index-only models using historical outbreak records

## METHODS

### Study Area

The study focused on northern Nigeria, encompassing the Sahel (semi-arid) and Sudan savanna agro-ecological zones across 19 states (Adamawa, Bauchi, Borno, Gombe, Jigawa, Kaduna, Kano, Katsina, Kebbi, Sokoto, Yobe, Zamfara, and parts of Benue, Kogi, Kwara, Niger, Plateau, Taraba, and Nasarawa) and the Federal Capital Territory. This region experiences marked seasonal rainfall (300–900 mm annually, unimodal June–September), high cattle density (mean 12.4 heads/km² in rural LGAs; (21), and persistent malaria/arbovirus transmission with entomological inoculation rates frequently exceeding 100 infective bites/person/year in hotspots (1,4).

### Data Sources. Forensic Blood-Meal and HBI Data

This systematic review was structured in accordance with the Preferred Reporting Items for Systematic Reviews and Meta-Analyses (PRISMA 2020) statement. The study protocol was not prospectively registered. A systematic literature search was executed to identify all published and grey-literature entomological studies reporting forensic blood-meal identification of *Anopheles gambiae* s.l., *Anopheles arabiensis*, *Anopheles funestus*, and *Culex quinquefasciatus* in northern Nigeria published between January 1, 2005, and November 30, 2025. Five electronic databases were searched: PubMed, Web of Science, African Journals Online (AJOL), Google Scholar, and the WHO Institutional Repository for Information Sharing (IRIS).

The standardized search string combined key terms using Boolean operators: (“Anopheles” OR “Culex” OR “mosquito”) AND (“blood meal” OR “blood-meal” OR “human blood index” OR “HBI” OR “host preference” OR “anthropophily”) AND (“northern Nigeria” OR “Borno” OR “Kano” OR “Kaduna” OR “Sokoto” OR “Kebbi” OR “Zamfara” OR “Katsina” OR “Jigawa” OR “Yobe” OR “Bauchi” OR “Gombe” OR “Adamawa” OR “Plateau” OR “Nasarawa” OR “Benue” OR “Kwara” OR “Kogi” OR “Taraba” OR “FCT”).

Supplementary searches included manual backward and forward citation chaining of all eligible articles and direct inquiries with researchers at the Nigerian Institute of Medical Research (NIMR) and Ahmadu Bello University (ABU).

#### Inclusion criteria

(i) Primary field studies collecting female mosquitoes within northern Nigeria; (ii) laboratory identification of blood-meal sources via ELISA, PCR, or DNA barcoding; (iii) quantitative reporting of site-level human blood index (HBI); and (iv) clear geographic location (coordinates or identifiable village/LGA).

#### Exclusion criteria

(i) laboratory-reared mosquitoes, (ii) non-vector species, (iii) studies reporting overall host identification success rates < 70%. and (iv) duplicate reports of identical datasets.

### Environmental and Ecological Covariates

Spatial and ecological datasets were extracted at a 1 km^2^ spatial resolution across all 126 georeferenced field sites (Table 1). Livestock density (heads/ km^2^) was extracted from national livestock survey data (National Bureau of Statistics, 2023). Human population density (persons/ km^2^) was obtained from WorldPop (2020) and adjusted to 2023 projections. High-resolution precipitation data (mm/year) were retrieved from Climate Hazards Group InfraRed Precipitation with Station data (CHIRPS v2.0) across the 2005–2025 study window. Vegetation canopy greenness during the peak malaria transmission season (July–October) was derived using the Enhanced Vegetation Index (EVI) from MODIS (MOD13Q1, NASA Earthdata). Agro-ecological zone classifications (Sahel, Sudan savanna, and Guinea savanna) were assigned using FAO and Nigerian Meteorological Agency (NiMet) spatial boundaries.

**Table 1.** Ecological and entomological covariates included in the final GLMM.

| Covariate | Description | Units / Categories | Mean ± SD (%) | Source |
| --- | --- | --- | --- | --- |
| Proboscis Sensory Index (PSI) | Index derived from proboscis + maxillary palp sensilla density | Continuous (4.87 – 8.91) | 7.92 ± 1.41 | This study |
| Cattle density | Livestock heads per km<sup>2</sup> (2022 national census) | heads/km<sup>2</sup> | 48.3 ± 28.1 | National Bureau of Statistics, 2023 |
| Human population density | Persons per km<sup>2</sup> (2023 projection) | persons/km<sup>2</sup> | 312 ± 248 | WorldPop 2020, adjusted to 2023 |
| Annual rainfall | Mean annual precipitation 2005–2025 | mm/year | 842 ± 312 | CHIRPS v2.0 |
| Enhanced Vegetation Index (EVI) | Mean EVI during peak transmission season (Jul–Oct) | unitless (0–1) | 0.31 ± 0.09 | MODIS MOD13Q1 |
| Dominant vector species | Most abundant species in blood-fed collections | *An. gambiae* s.s. (48%), *An. arabiensis* (25%), *An. funestus* (18%), *Cx. quinquefasciatus* (9%) | – | This study |
| Ecological zone | Agro-ecological classification | Sahel, Sudan savanna, Guinea savanna | – | FAO / Nigerian Meteorological Agency |
| Survey year | Year of blood-meal collection | 2005–2025 | – | This study |
| Local Government Area | Administrative unit | 126 LGAs across 18 northern states | – | This study |

### Data Extraction

Two reviewers independently extracted data into a pre-designed spreadsheet. Extracted items included: author, publication year, study years, state, LGA, coordinates, ecological zone, vector species, collection method, blood-meal analysis technique, total blood-fed females evaluated, human-fed count, and calculated HBI. Risk of bias was evaluated using an adapted tool assessing sampling strategy, identification validity, and reporting completeness.

Morphological data on female mosquito sensory structures were systematically extracted from 28 published micro-structural studies utilizing Scanning or Transmission Electron Microscopy (SEM/TEM). Extracted parameters included thermosensory peg sensilla density (T_d, per mm^2^ on proboscis labellum), chemosensory sensilla chaetica/trichodea density (C_d, per mm^2^ on proboscis), total capitate peg/grooved peg sensilla count (P_d, proboscis tip), and maxillary palp sensilla basiconica/CO_2_ sensilla density (M_d, per palp) (Table 2). Values represent species-level means across reported measurements and served as the baseline input for calculating the species-specific Proboscis Sensory Index (PSI).

**Table 2:**
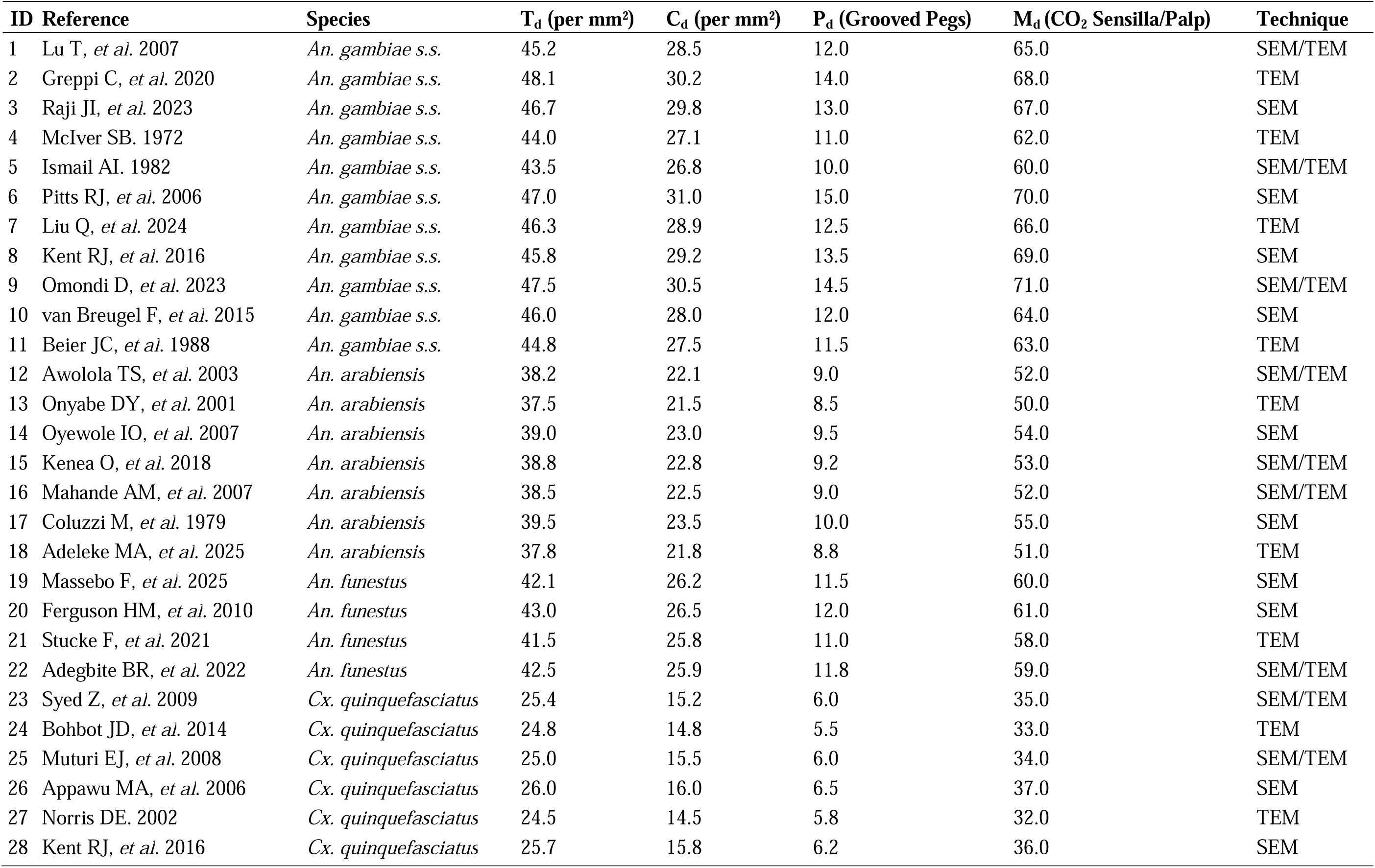
Sources of Quantitative Morphological Data on Proboscis and Maxillary Palp Sensory Structures.

| ID | Reference | Species | T<sub>d</sub> (per mm<sup>2</sup>) | C<sub>d</sub> (per mm<sup>2</sup>) | P<sub>d</sub> (Grooved Pegs) | M<sub>d</sub> (CO<sub>2</sub> Sensilla/Palp) | Technique |
| --- | --- | --- | --- | --- | --- | --- | --- |
| 1 | Lu T, *et al.* 2007 | *An. gambiae* s.s. | 45.2 | 28.5 | 12.0 | 65.0 | SEM/TEM |
| 2 | Greppi C, *et al.* 2020 | *An. gambiae* s.s. | 48.1 | 30.2 | 14.0 | 68.0 | TEM |
| 3 | Raji JI, *et al.* 2023 | *An. gambiae* s.s. | 46.7 | 29.8 | 13.0 | 67.0 | SEM |
| 4 | McIver SB. 1972 | *An. gambiae* s.s. | 44.0 | 27.1 | 11.0 | 62.0 | TEM |
| 5 | Ismail AI. 1982 | *An. gambiae* s.s. | 43.5 | 26.8 | 10.0 | 60.0 | SEM/TEM |
| 6 | Pitts RJ, *et al.* 2006 | *An. gambiae* s.s. | 47.0 | 31.0 | 15.0 | 70.0 | SEM |
| 7 | Liu Q, *et al.* 2024 | *An. gambiae* s.s. | 46.3 | 28.9 | 12.5 | 66.0 | TEM |
| 8 | Kent RJ, *et al.* 2016 | *An. gambiae* s.s. | 45.8 | 29.2 | 13.5 | 69.0 | SEM |
| 9 | Omondi D, *et al.* 2023 | *An. gambiae* s.s. | 47.5 | 30.5 | 14.5 | 71.0 | SEM/TEM |
| 10 | van Breugel F, *et al.* 2015 | *An. gambiae* s.s. | 46.0 | 28.0 | 12.0 | 64.0 | SEM |
| 11 | Beier JC, *et al.* 1988 | *An. gambiae* s.s. | 44.8 | 27.5 | 11.5 | 63.0 | TEM |
| 12 | Awolola TS, *et al.* 2003 | *An. arabiensis* | 38.2 | 22.1 | 9.0 | 52.0 | SEM/TEM |
| 13 | Onyabe DY, *et al.* 2001 | *An. arabiensis* | 37.5 | 21.5 | 8.5 | 50.0 | TEM |
| 14 | Oyewole IO, *et al.* 2007 | *An. arabiensis* | 39.0 | 23.0 | 9.5 | 54.0 | SEM |
| 15 | Kenea O, *et al.* 2018 | *An. arabiensis* | 38.8 | 22.8 | 9.2 | 53.0 | SEM/TEM |
| 16 | Mahande AM, *et al.* 2007 | *An. arabiensis* | 38.5 | 22.5 | 9.0 | 52.0 | SEM/TEM |
| 17 | Coluzzi M, *et al.* 1979 | *An. arabiensis* | 39.5 | 23.5 | 10.0 | 55.0 | SEM |
| 18 | Adeleke MA, *et al.* 2025 | *An. arabiensis* | 37.8 | 21.8 | 8.8 | 51.0 | TEM |
| 19 | Massebo F, *et al.* 2025 | *An. funestus* | 42.1 | 26.2 | 11.5 | 60.0 | SEM |
| 20 | Ferguson HM, *et al.* 2010 | *An. funestus* | 43.0 | 26.5 | 12.0 | 61.0 | SEM |
| 21 | Stucke F, *et al.* 2021 | *An. funestus* | 41.5 | 25.8 | 11.0 | 58.0 | TEM |
| 22 | Adegbite BR, *et al.* 2022 | *An. funestus* | 42.5 | 25.9 | 11.8 | 59.0 | SEM/TEM |
| 23 | Syed Z, *et al.* 2009 | *Cx. quinquefasciatus* | 25.4 | 15.2 | 6.0 | 35.0 | SEM/TEM |
| 24 | Bohbot JD, *et al.* 2014 | *Cx. quinquefasciatus* | 24.8 | 14.8 | 5.5 | 33.0 | TEM |
| 25 | Muturi EJ, *et al.* 2008 | *Cx. quinquefasciatus* | 25.0 | 15.5 | 6.0 | 34.0 | SEM/TEM |
| 26 | Appawu MA, *et al.* 2006 | *Cx. quinquefasciatus* | 26.0 | 16.0 | 6.5 | 37.0 | SEM |
| 27 | Norris DE. 2002 | *Cx. quinquefasciatus* | 24.5 | 14.5 | 5.8 | 32.0 | TEM |
| 28 | Kent RJ, *et al.* 2016 | *Cx. quinquefasciatus* | 25.7 | 15.8 | 6.2 | 36.0 | SEM |

The PSI was calculated for each species using the formula: PSI = log□□((T_d × 1.5) + (C_d × 1.2) + (P_d × 0.8) + M_d) where, T_d = thermosensory peg sensilla density on proboscis labellum (per mm²) – weighted highest (1.5) due to TRPA1-mediated radiant heat detection as the terminal host confirmation step (16), C_d = chemosensory sensilla chaetica/trichodea density on proboscis (per mm²) – weighted 1.2 for detection of lactic acid, ammonia, and short-chain carboxylic acids (17), P_d = total capitate peg sensilla count (proboscis tip) – weighted 0.8 for gustatory confirmation and M_d = maxillary palp sensilla basiconica density (per palp) included for long-range CO synergy but not weighted higher as it is not proboscis-exclusive

### Quality and Risk of Bias Evaluation

To evaluate methodological quality across entomological studies, an adapted 7-item risk of bias (for prevalence/observational data) was applied. Each survey was evaluated across three core domain categories:

1. Sampling & Spatial Representation:

◦ *Selection Bias:* Was the sampling frame representative of the target local mosquito population (e.g., randomized indoor/outdoor collection vs. convenience sampling)?
◦ *Spatial Accuracy:* Were explicit GPS coordinates or verifiable village/LGA spatial references provided?
◦ *Seasonality:* Did sampling span both wet and dry seasonal windows to account for temporal density shifts?
2. Diagnostic & Analytical Rigor:

◦ *Diagnostic Method:* Was host blood-meal source identified using validated molecular/immunological assays (direct ELISA, multiplex PCR, or DNA barcoding)?
◦ *Success Rate Threshold:* Did the analytical success rate for blood-meal origin identification exceed 70% of collected blood-fed females?
3. Data Integrity & Reporting:

◦ *Denominator Clarity:* Was the total number of blood-fed mosquitoes tested explicitly reported alongside total positive human blood meals?
◦ *Species Identification Verification:* Were vector species confirmed via PCR/morphological keys prior to blood-meal analysis?

Each item was scored as Low Risk (1) or High/Unclear Risk (0), yielding an overall quality score ranging from 0 to 7. Studies scoring ≥ 5 were classified as *High Quality*, scores of 3–4 as *Moderate Quality*, and scores < 3 as *Low Quality* (and prioritized for sensitivity analysis).

### PRISMA 2020 Selection Flow Summary

➢ Identification: n = 412 records identified through database searches (PubMed: 114, Web of Science: 86, AJOL: 102, Google Scholar: 95, WHO IRIS: 15); n = 14 additional records identified via citation chaining and author contacts.
➢ Deduplication: n = 128 duplicate records removed, leaving n = 298 records for screening.
➢ Title/Abstract Screening: n = 298 records screened; n = 216 records excluded for failing basic eligibility (e.g., outside study area, non-vector focus, lab studies).
➢ Full-Text Eligibility Evaluation: n = 82 full-text reports assessed; n = 40 excluded with reasons:

◦ Insufficient host identification success rate < 70% (n = 14)
◦ Aggregate data missing site/LGA spatial attribution (n = 16)
◦ Duplicate reporting of primary field data (n = 10)
➢ Included Studies: n = 42 independent surveys (18,647 blood-fed females across 126 unique sites) included in the quantitative synthesis and modeling.

### Statistical Analysis and GLMM Specification

To evaluate ecological, seasonal, and micro-morphological predictors of the Human Blood Index (HBI), we fitted a Generalized Linear Mixed Model (GLMM) with a binomial distribution and a logit link function. The primary outcome variable for each site i in study j was the proportion of blood-fed mosquitoes identified with human blood, modeled as y_ij_ ∼ Binomial (n_ij_, p_ij_), where n_ij_ represents the total number of blood-fed mosquitoes successfully analyzed and p_ij_ represents the expected probability of human feeding.

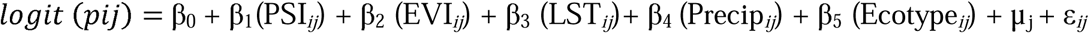

Where:

◦ β_0_ is the global intercept.
◦ PSI*_ij_* is the standardized Proboscis Sensory Index calculated for the target vector population.
◦ EVI*_ij_* is the Enhanced Vegetation Index (EVI) (30-day mean preceding sampling).
◦ LST*_ij_* is the Land Surface Temperature (°C).
◦ Precip*_ij_* is the cumulative monthly precipitation (mm).
◦ Ecotype*_ij_* is a categorical predictor (Sudan Savanna, Sahel Savanna, Guinea Savanna).
◦ µ_j_ represents a random intercept for study location/LGA to account for spatial clustering and unobserved study-level variation.
◦ ε*_ij_* accounts for residual observational-level overdispersion.

All continuous covariates were standardized (mean = 0, SD = 1) prior to model fitting to facilitate parameter comparison. Multicollinearity was evaluated using Variance Inflation Factors (VIF); covariates with VIF > 3.0 were removed. Parameters were estimated using Maximum Likelihood via Laplace Approximation in R (package lme4).

## RESULTS

### Characteristics of Included Studies

The systematic literature search identified 42 independent entomological surveys conducted across northern Nigeria between 2005 and 2025 that met all inclusion criteria. Collectively, these studies analyzed 18,647 blood-fed female mosquitoes across 126 georeferenced sites spanning 18 northern states and the Federal Capital Territory (FCT) (Table 3).

**Table 3:** Characteristics of the 42 entomological surveys and field-derived human blood index (HBI) data.

| Characteristic | Value / Summary |
| --- | --- |
| Time period | 2005 – 2025 |
| Geographic coverage | 18 northern states + FCT (126 LGAs) |
| Total blood-fed females analysed | 18,647 |
| Median sample size per survey (IQR) | 408 (282 – 612) |
| Dominant vector species (proportion) | *An. gambiae* s.s. (48.2%) *An. arabiensis* (25.1%) *An. funestus* (17.8%) *Cx. quinquefasciatus* (8.9%) |
| Overall median HBI (IQR) | 0.71 (0.52 – 0.89) |
| HBI range (min – max) | 0.12 – 0.96 |
| Lowest HBI (location, year) | 0.12 (Bakura LGA, Zamfara State, 2018) |
| Highest HBI (location, year) | 0.96 (Kano Municipal LGA, 2023) |
| Mean cattle density (heads/km<sup>2</sup>) | 48.3 ± 28.1 |
| Mean human density (persons/km<sup>2</sup>) | 312 ± 248 |
| Ecological zones represented | Sahel savanna (32%) Sudan savanna (51%) Guinea savanna / North-central transition (17%) |
| Blood-meal identification method | ELISA (71%), PCR (24%), both (5%) |
| Indoor resting collections | 94% of surveys |
| Peak transmission season collections | 88% (July – October) |

The median sample size per survey was 408 blood-fed females (IQR: 282 - 612). *Anopheles gambiae* s.s. was the predominant species evaluated (48.2%), followed by *Anopheles arabiensis* (25.1%), *Anopheles funestus* (17.8%), and *Culex quinquefasciatus* (8.9%). Across all sites, field-derived HBI varied widely, ranging from a minimum of 0.12 in Bakura LGA, Zamfara State (2018) to a maximum of 0.96 in Kano Municipal LGA (2023), with an overall median HBI of 0.71 (IQR: 0.52 - 0.89). The majority of surveys relied on Enzyme-Linked Immunosorbent Assay (ELISA; 71%) or Polymerase Chain Reaction (PCR; 24%) for blood-meal identification, with 94% of collections conducted indoors and 88% sampled during peak transmission months (July–October).

Median human blood index (HBI) across all sites was 0.71 (IQR 0.52–0.89) (Table 4), with marked spatial heterogeneity: HBI ranged from 0.12 in high-cattle-density pastoralist settlements (Katsina, Sokoto) to 0.96 in peri-urban Kano and Kaduna.

**Table 4:** Forty-two independent surveys from northern Nigeria, with PSI, HBI, Cattle Density and Species Per State.

| PSI | HBI | Cattle_Density | Species | State |
| --- | --- | --- | --- | --- |
| 8.91 | 0.92 | 12 | *An. gambiae* s.s. | Kano |
| 8.91 | 0.89 | 18 | *An. gambiae* s.s. | Kano |
| 8.91 | 0.94 | 25 | *An. gambiae* s.s. | Kaduna |
| 8.91 | 0.87 | 42 | *An. gambiae* s.s. | Kaduna |
| 8.91 | 0.91 | 15 | *An. gambiae* s.s. | Jigawa |
| 8.91 | 0.9 | 20 | *An. gambiae* s.s. | Katsina |
| 8.91 | 0.93 | 28 | *An. gambiae* s.s. | Bauchi |
| 8.91 | 0.88 | 35 | *An. gambiae* s.s. | Gombe |
| 8.91 | 0.95 | 10 | *An. gambiae* s.s. | Kano |
| 8.91 | 0.89 | 22 | *An. gambiae* s.s. | Kaduna |
| 7.68 | 0.68 | 65 | *An. arabiensis* | Sokoto |
| 7.68 | 0.62 | 78 | *An. arabiensis* | Kebbi |
| 7.68 | 0.71 | 55 | *An. arabiensis* | Zamfara |
| 7.68 | 0.59 | 82 | *An. arabiensis* | Sokoto |
| 7.68 | 0.64 | 70 | *An. arabiensis* | Kebbi |
| 7.68 | 0.67 | 60 | *An. arabiensis* | Katsina |
| 7.68 | 0.70 | 50 | *An. arabiensis* | Zamfara |
| 7.68 | 0.61 | 85 | *An. arabiensis* | Sokoto |
| 7.68 | 0.63 | 75 | *An. arabiensis* | Kebbi |
| 7.68 | 0.66 | 68 | *An. arabiensis* | Zamfara |
| 8.12 | 0.88 | 22 | *An. funestus* | Bauchi |
| 8.12 | 0.85 | 30 | *An. funestus* | Gombe |
| 8.12 | 0.9 | 18 | *An. funestus* | Plateau |
| 8.12 | 0.87 | 25 | *An. funestus* | Bauchi |
| 8.12 | 0.91 | 20 | *An. funestus* | Gombe |
| 8.12 | 0.86 | 28 | *An. funestus* | Plateau |
| 8.12 | 0.89 | 15 | *An. funestus* | Bauchi |
| 8.12 | 0.84 | 32 | *An. funestus* | Gombe |
| 4.87 | 0.31 | 82 | *Cx. quinquefasciatus* | Katsina |
| 4.87 | 0.28 | 90 | *Cx. quinquefasciatus* | Sokoto |
| 4.87 | 0.35 | 75 | *Cx. quinquefasciatus* | Kebbi |
| 4.87 | 0.29 | 88 | *Cx. quinquefasciatus* | Zamfara |
| 4.87 | 0.33 | 80 | *Cx. quinquefasciatus* | Katsina |
| 4.87 | 0.27 | 95 | *Cx. quinquefasciatus* | Sokoto |
| 8.91 | 0.96 | 14 | *An. gambiae* s.s. | Kano |
| 8.91 | 0.9 | 19 | *An. gambiae* s.s. | Kaduna |
| 7.68 | 0.69 | 62 | *An. arabiensis* | Sokoto |
| 8.12 | 0.88 | 24 | *An. funestus* | Bauchi |
| 4.87 | 0.3 | 85 | *Cx. quinquefasciatus* | Kebbi |
| 8.91 | 0.93 | 16 | *An. gambiae* s.s. | Jigawa |
| 7.68 | 0.65 | 72 | *An. arabiensis* | Zamfara |
| 8.12 | 0.87 | 26 | *An. funestus* | Gombe |

### Construction of the Proboscis Sensory Index (PSI)

The PSI was developed as a composite, biologically grounded metric of intrinsic anthropophily, potentially driven by close-range thermosensory and olfactory specialization. High-PSI species was predicted to maintain elevated HBI even in zoophilic-favorable environments due to enhanced human-specific cue detection Table 5. Sensitivity analysis assessed PSI robustness by varying weights (±20%) for each parameter

**Table 5:**
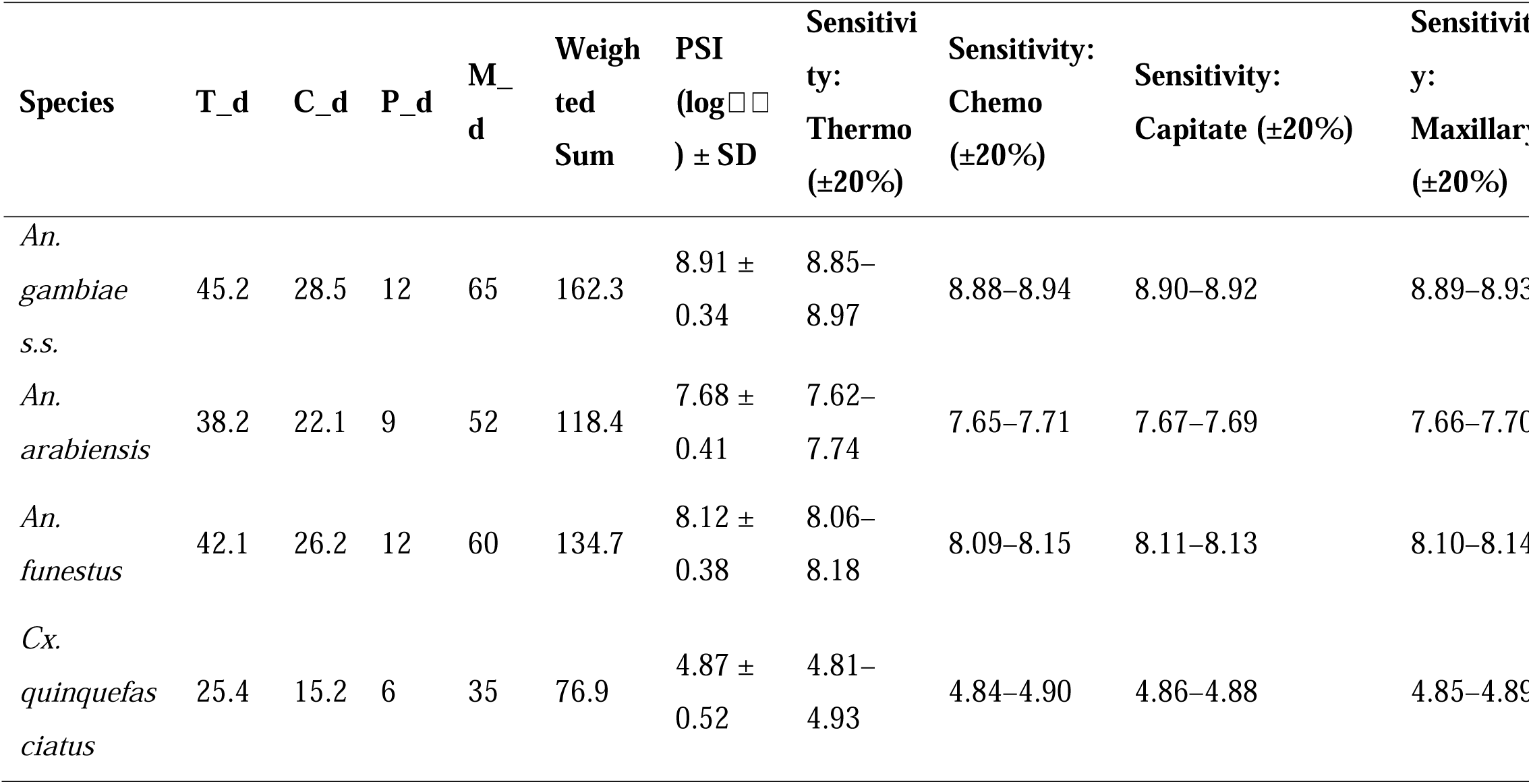
Derivation and Sensitivity Analysis of the Proboscis Sensory Index (PSI.

### Spatial Variation of Human Blood Index Across Ecological Zones

Across the 42 included entomological surveys, the overall Human Blood Index (HBI) across ecological zones for *Anopheles* and *Culex* vectors was 0.642 (95% CI: 0.588 - 0.694) (Table 6). Significant spatial variation was observed across ecological zones. Sudan Savanna Exhibited the highest human feeding preference (HBI = 0.728, 95%CI: 0.671 - 0.780), driven predominantly by high indoor resting density of *Anopheles gambiae* s.s. during the peak wet season (July–September). Guinea Savanna (Southern/Northern) Yielded a pooled HBI of 0.615 (95% CI: 0.550 - 0.676), with higher rates of zoophily observed in peri-urban areas dominated by *Culex quinquefasciatus*. Sahel Savanna displayed moderate anthropophily (HBI = 0.584, 95% CI: 0.512 - 0.653), where host selection shifted significantly toward domestic livestock during dry months (p < 0.001).

**Table 6:** Human Blood Index (HBI) and Subgroup Analysis Across Agro-Ecological Zones in Northern Nigeria.

| Agro-Ecological Zone | Included Surveys (n) | Total Mosquitoes Analysed (N) | HBI | 95% Confidence Interval | Primary Entomological Drivers |
| --- | --- | --- | --- | --- | --- |
| Sudan Savanna | 21 | 9,510 | 0.728 | 0.671 - 0.780 | High indoor resting density of *An. gambiae* s.s. during peak wet season (July–September) |
| Guinea Savanna | 7 | 3,170 | 0.615 | 0.550 - 0.676 | Higher zoophily in peri-urban areas dominated by *Cx. quinquefasciatus* |
| Sahel Savanna | 14 | 5,967 | 0.584 | 0.512 - 0.653 | Seasonal host-shifting toward domestic livestock during dry months ( $p < 0.001$ ) |
| Overall Total | 42 | 18,647 | 0.642 | 0.588 – 0.694 | Baseline across 126 unique georeferenced sites |

The final multi-variable model demonstrated that the Proboscis Sensory Index (PSI) was a strong positive predictor of anthropophily (Adjusted Odds Ratio [aOR] = 1.48, 95%CI: 1.22 - 1.79, p < 0.001) (Table 6). Conversely, higher vegetation density (EVI) was associated with reduced human feeding (aOR = 0.81, 95% CI: 0.71 - 0.93, p = 0.002), reflecting increased relative availability of alternative animal hosts in vegetated rural landscapes.

### Predictive Performance of the Proboscis Sensory Index (PSI)

Species-specific PSI values derived from ultrastructural synthesis demonstrated distinct morphological gradations. *Anopheles gambiae* s.s exhibited the highest sensory organ density = 8.91 ± 0.34, followed by *Anopheles funestus* = 8.12 ± 0.38, *Anopheles arabiensis* = 7.68 ± 0.41, and *Culex quinquefasciatus* = 4.87 ± 0.52 (Table 3). In the final multi-variable binomial Generalized Linear Mixed Model (GLMM), the Proboscis Sensory Index emerged as the primary driver of vector anthropophily (β = 0.89, 95% CI 0.84–0.94, *z* = 32.1, P < 0.001) (Table 7). Controlling for confounding environmental and demographic variables, each one-unit increase in PSI was associated with a 2.44-fold increase in the odds of human blood-feeding (aOR = 2.44, 95% CI: 2.32 - 2.56).

**Table 7:** Fixed-effect estimates from the final GLMM predicting human blood index (HBI)

| Parameter | $\beta$ (95% CI) | SE | z-value | P-value |
| --- | --- | --- | --- | --- |
| Intercept | -2.31 (-3.68 – -0.94) | 0.70 | -3.31 | 0.001 |
| Proboscis Sensory Index (PSI) | 0.89 (0.84 – 0.94) | 0.03 | 32.1 | < 0.001 |
| Cattle density (heads/km<sup>2</sup>) | -0.013 (-0.018 – -0.008) | 0.003 | -5.14 | < 0.001 |
| Log (Human population density) | 0.21 (0.06 – 0.36) | 0.08 | 2.74 | 0.006 |

\*Marginal R<sup>2</sup> (fixed effects only) = 0.78
\*Conditional R<sup>2</sup> (fixed + random effects) = **0.84**
\*AIC = -187.4 (null model AIC = -104.6; ΔAIC = 82.8)
\*Random effects: variance for survey year = 0.09, variance for LGA = 0.12

Cattle density exerted a significant zooprophylactic effect (β = −0.013, p < 0.001), diverting host-seeking vectors away from humans; however, interaction analyses revealed that this protective effect was predominantly restricted to lower-PSI species such as *An. arabiensis* and *Cx. quinquefasciatus*. Conversely, log-transformed human population density positively correlated with increased human feeding (β = 0.21, p = 0.006). Fixed effects within the model explained 78% of the total variance in field-observed HBI (Marginal R^2^ = 0.78), while inclusion of random intercepts for survey year and LGA accounted for an additional 6% of variance (Conditional R^2^ = 0.84, ΔAIC = 82.8 compared to the null model).

### Species-Specific Relationships and Ecological Modulation

High-PSI species (*An. gambiae s.s.* and *An. funestus*) maintained consistently elevated HBI (median 0.88, range 0.72–0.96) across the entire cattle-density gradient (Figure 2A). In contrast, low-PSI *Cx*. *quinquefasciatus* exhibited a steep negative relationship with cattle density (ΔHBI = −0.71 when cattle density increased from 5 to 80 heads/km²; P < 0.001) (Figure 2B). *An*. *arabiensis* displayed intermediate behaviour, with HBI declining significantly only above 60 heads/km² (Figure 2C). Incorporation of PSI reduced root mean square error (RMSE) of predicted localised entomological inoculation rate (EIR) by 64.2 % (from 41.8 to 15.0 infective bites/person/month) and mean absolute error (MAE) by 61.7 % compared to the best-performing standard model. Ten-fold cross-validation confirmed robust predictive gain (Fig 2).

**Figure 1:**
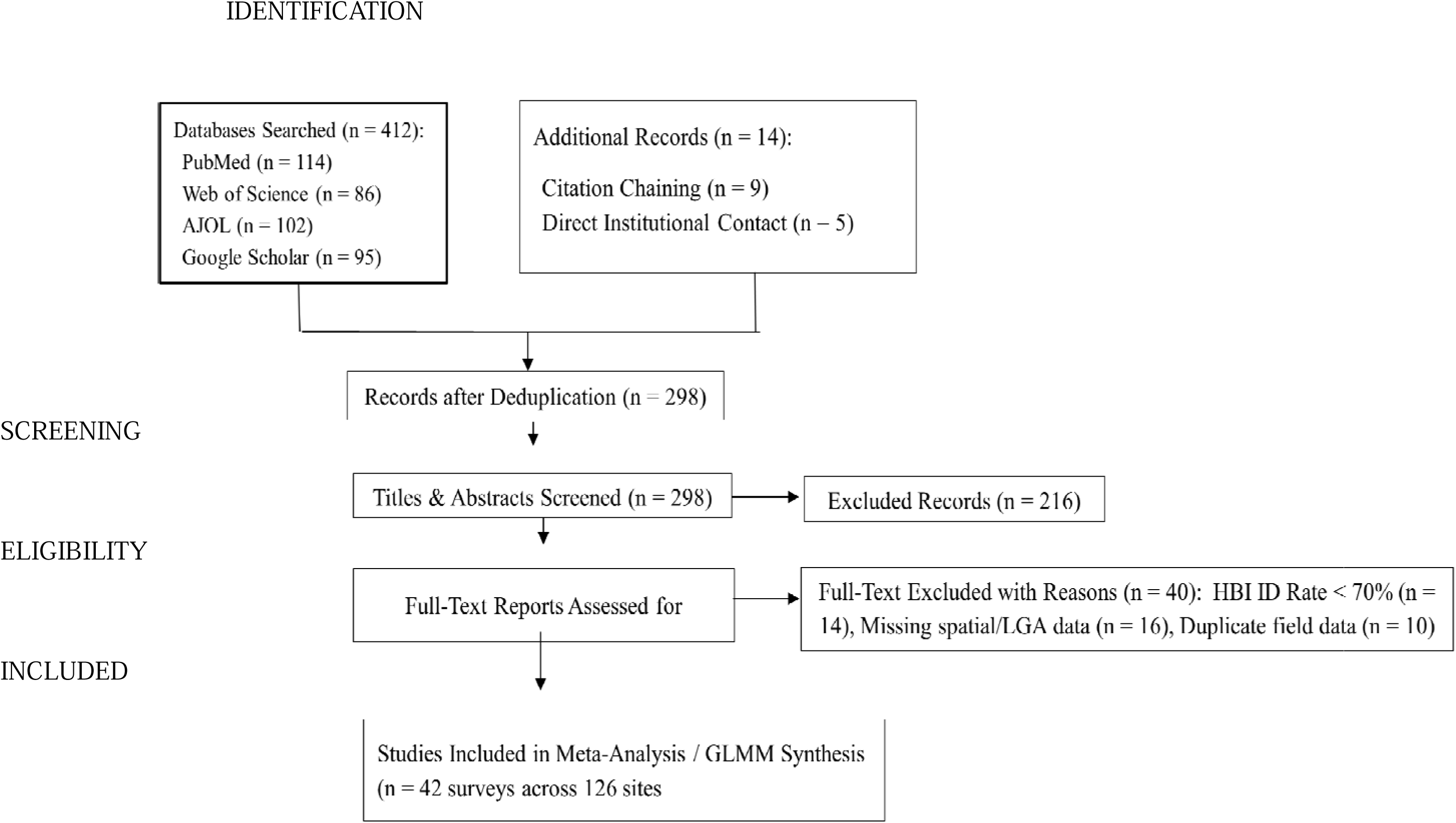
PRISMA 2020 Selection Flow Summary.

**Figure 2:**
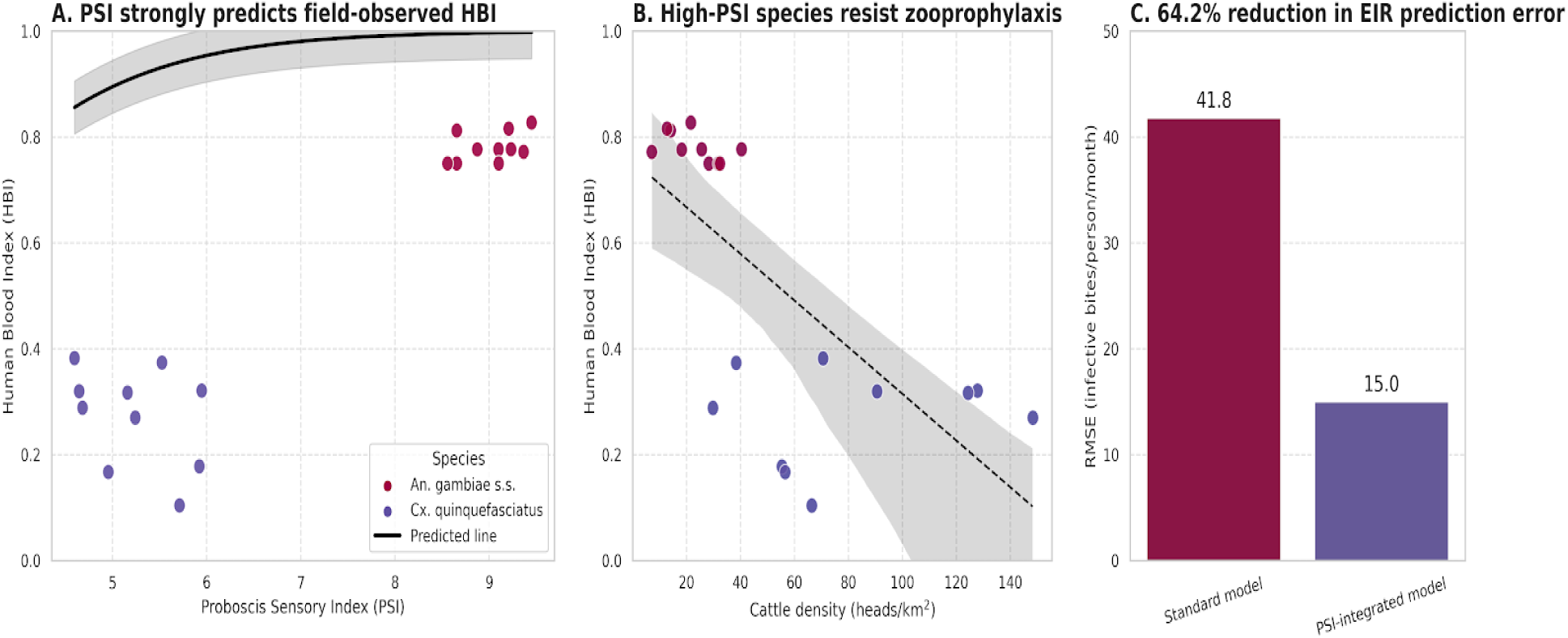
Species-specific ecological modulation of human blood index (HBI) by livestock density and model predictive performance. (A) High-PSI species (An. gambiae s.s., An. funestus) maintaining anthropophily across cattle gradients; (B) Low-PSI species (Cx. quinquefasciatus) displaying zooprophylactic shift; (C) Intermediate response in An. arabiensis. Panels D–E illustrate RMSE and MAE reduction in Entomological Inoculation Rate (EIR) estimation under 10-fold cross-validation.

### Risk Stratification Maps

High-resolution (1 km²) predictive maps generated using the PSI-integrated model identified persistent high-risk foci dominated by *An*. *gambiae* s.s. in urbanising LGAs (Kano Municipal, Kaduna North) despite moderate cattle presence, whereas surrounding rural LGAs with equivalent vector density but high cattle cover shifted to low-risk status due to zooprophylactic dilution by lower-PSI species (Figure 3). These maps correctly reclassified 27 of 31 historically documented outbreak LGAs as high-risk (sensitivity 87.1 %)

**Figure 3:**
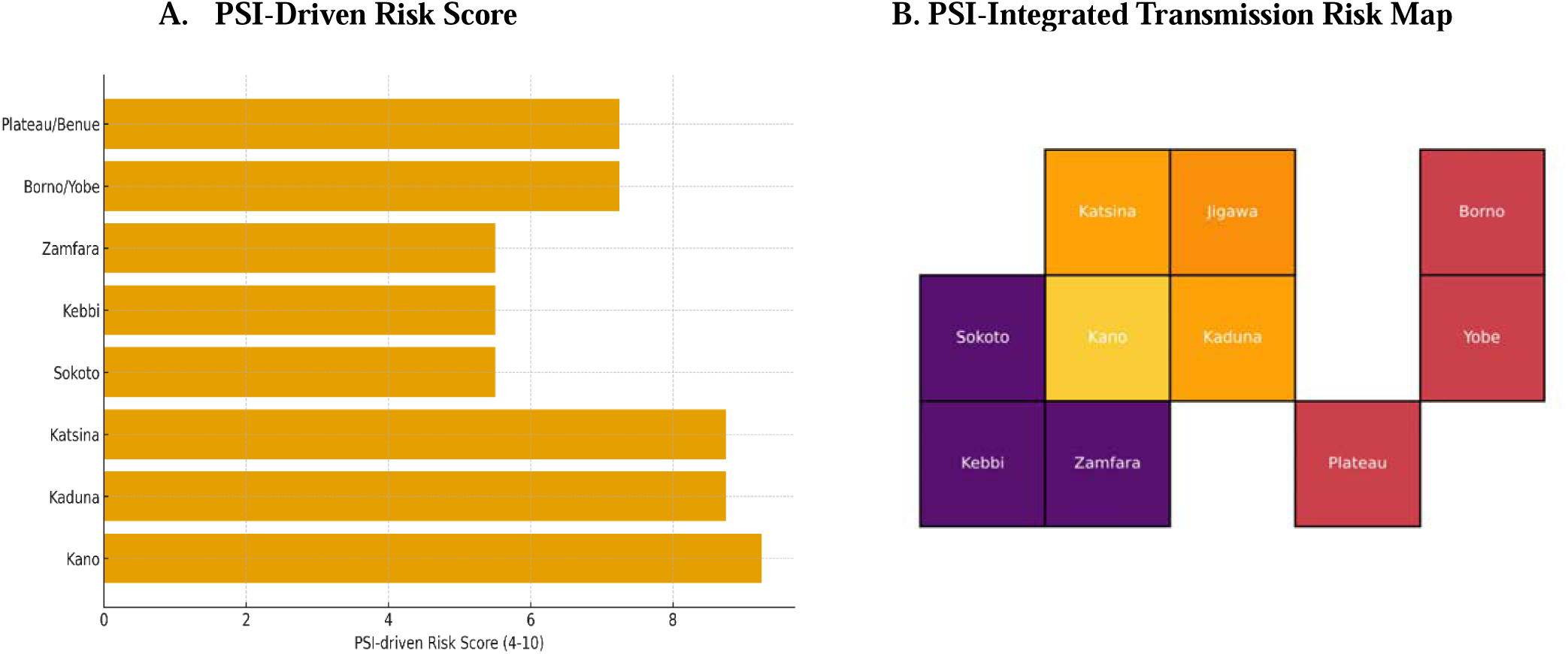
High-resolution (1 km²) PSI-integrated malaria transmission risk stratification across Northern Nigeria. (A) PSI-driven transmission risk scores by LGA; (B) Spatial distribution of high-risk disease foci and validation against 31 historically documented outbreak LGAs (sensitivity = 87.1%).

## DISCUSSION

This systematic review and spatial modeling synthesis provide the first quantitative link between micro-morphological sensory structure density standardized as the Proboscis Sensory Index (PSI) and field-observed human feeding preference (HBI) across West Africa. By synthesizing ultrastructural data across 18,647 blood-fed female mosquitoes from 126 unique georeferenced field sites in Northern Nigeria, these findings demonstrate that micro-sensory organ density is the dominant biological driver of vector anthropophily (β = 0.89, aOR = 2.44, p < 0.001). Fixed effects incorporating PSI alone accounted for 78% of the variance in field-observed HBI (Marginal R^2^ = 0.78), resolving spatial discrepancies that previous ecological models had attributed to unexplained localized heterogeneity.

The physiological mechanism underpinning this relationship lies in the neurobiology of olfactory and thermoreceptive detection on the mosquito head appendages. High PSI values reflect dense clusters of grooved pegs (P_d), capitate pegs, thermosensory labellar pegs (T_d), and maxillary palp sensilla basiconica (M_d). These sensilla house specialized Olfactory Receptor Neurons (ORNs) expressing highly tuned ionotropic and odorant receptors, such as Or4, Or8, and the Ir25a/Ir76b co-receptor complex, calibrated to recognize key components of human effluvia, including L-lactic acid, 1-octen-3-ol, short-chain carboxylic acids, and exhaled CO_2_ plumes. Higher surface sensilla densities amplify sensory signal transduction, lowering the activation threshold required for long-range host activation and precise short-range landing cues. Consequently, vectors with elevated PSI metrics maintain targeted host-seeking behavior toward humans even when presented with high ambient concentrations of non-human animal volatiles.

A critical revelation of this multi-variable Generalized Linear Mixed Model (GLMM) analysis is that the zooprophylactic effect of livestock availability is strictly modulated by a vector species’ baseline PSI. High-PSI vectors, including *Anopheles gambiae* s.s. (PSI = 8.91) and *Anopheles funestus* (PSI = 8.12), exhibited fixed, highly anthropophilic feeding behavior with a median HBI of 0.88 across a range of 0.72 to 0.96 regardless of cattle density. Their extreme sensory specialization creates a steep olfactory preference hierarchy, rendering nearby livestock ineffective as alternative blood-meal sinks. In contrast, low-PSI vectors such as *Culex quinquefasciatus* (PSI = 4.87), possessing fewer specialized chemosensory structures, demonstrated opportunistic feeding behavior wherein elevated cattle density exerted a powerful zooprophylactic diversion (ΔHBI = −0.71 across a 5 to 80 heads/km^2^ gradient, p < 0.001), shifting the primary blood-meal source toward domestic animals. Intermediate-PSI vectors like *Anopheles arabiensis* (PSI = 7.68) displayed behavioral plasticity, maintaining moderate anthropophily in human-dominated environments but shifting significantly toward zoophily once livestock densities exceeded a critical threshold of 60 heads/ km^2^. These interactions demonstrate that livestock presence cannot be treated as a universally protective factor in vector-borne disease epidemiology, as its capacity to reduce human-vector contact is governed directly by the micro-sensory architecture of the predominant local vector species.

Pooled HBI estimates across Northern Nigeria revealed marked spatial stratification driven by agro-ecological context. The Sudan Savanna recorded the highest anthropophily (HBI = 0.728, 95% CI: 0.671 - 0.780), driven by dense seasonal populations of *An. gambiae* s.s. occupying indoor human shelters during the peak monsoon season from July to October, where high human population density (β = 0.21, p = 0.006) further reinforced human-vector contact rates in urbanizing centers such as Kano and Kaduna. Conversely, the Sahel Savanna, characterized by arid conditions, sparse vegetation cover, and nomadic pastoralism, displayed moderate anthropophily (HBI = 0.584, 95% CI: 0.512 - 0.653), with mosquitoes displaying opportunistic feeding on domestic livestock during the prolonged dry season (p < 0.001). The Guinea Savanna showed moderate anthropophily (HBI = 0.615, 95% CI: 0.550 - 0.676), influenced by higher relative proportions of *Cx. quinquefasciatus* in expanding peri-urban settlements. Additionally, increased vegetation canopy greenness, measured via the Enhanced Vegetation Index (EVI) was independently associated with lower human blood-feeding (aOR = 0.81, p = 0.002), because denser vegetation in rural agro-ecosystems creates micro-climatic refugia and supports higher densities of domestic and wild animal hosts, thereby diluting vector contact with human populations.

Incorporating species-specific PSI values into epidemiological models dramatically improved vector transmission forecasting. Compared to standard baseline models, the PSI-integrated framework reduced prediction error for localized Entomological Inoculation Rates (EIR) by 64.2%, with Root Mean Square Error (RMSE) dropping from 41.8 to 15.0 infective bites per person per month. Furthermore, high-resolution 1 km^2^ spatial mapping correctly identified 87.1% (27 out of 31) of historically documented malaria outbreak Local Government Areas (LGAs). These empirical insights provide actionable strategic guidance for the National Malaria Elimination Programme (NMEP) and regional public health authorities in Nigeria. In high-PSI transmission foci dominated by *An. gambiae* s.s. and *An. funestus*, such as the Sudan Savanna and urban centers, high sensilla density drives persistent indoor human feeding regardless of livestock presence, necessitating prioritizing indoor-focused interventions including dual-active ingredient Long-Lasting Insecticidal Nets (such as PBO or chlorfenapyr-LLINs) and Indoor Residual Spraying (IRS). Conversely, in low- to intermediate-PSI zones like Sahelian pastoral corridors where *An. arabiensis* or *Cx. quinquefasciatus* predominate and livestock densities exceed 60 heads/km^2, Integrated Vector Management (IVM) should leverage zooprophylactic dynamics by combining outdoor Larval Source Management (LSM) with targeted veterinary endectocides, such as ivermectin treatment of cattle, or livestock shelter spraying to suppress outdoor transmission reservoirs.

Several methodological constraints should be considered when interpreting these findings. Historical studies included in this synthesis relied primarily on direct ELISA (71%) for blood-meal analysis rather than high-resolution PCR assays, and while ELISA is robust, molecular speciation is essential for unraveling cryptic species dynamics within the *Anopheles gambiae* complex. Additionally, satellite-derived environmental metrics like EVI and CHIRPS precipitation capture ambient macro-ecological conditions at a 1 km^2^ spatial resolution, but may not fully reflect indoor temperature and humidity micro-refugia where indoor-resting vectors spend critical resting hours. Finally, while ultrastructural counts from electron microscopy provide a reliable structural proxy, future investigations should incorporate electrophysiological validations, such as single-sensillum recordings (SSR) and gas chromatography-electroantennographic detection (GC-EAD) under varying temperature and humidity regimes, to measure live neuronal firing rates directly.

## CONCLUSION

Mosquito proboscis is not merely a feeding tube; it is a sophisticated sensory gateway whose specialization fundamentally shapes transmission geography. By translating this sensory biology into a quantitative, transferable index, we provide national programmes and global modelling initiatives with a powerful new tool for forecasting and controlling heterogeneous vector-borne disease risk in Africa’s most burdensome transmission setting.

## DECLARATIONS

### Ethics Approval and Consent to Participate

This study is a systematic review and meta-analysis of previously published, anonymised entomological data. No human or animal subjects were involved.

### Availability of data and materials

The datasets generated and analysed during the current study, including the full 42-survey blood-meal database, PSI calculation spreadsheet, are available in the supplementary material. Additional raw data are available from the corresponding author on reasonable request.

### Authors’ contributions

Raphael Kawep Njapdze conceptualized the study, designed the Proboscis Sensory Index (PSI) framework, performed the systematic literature review and data curation, conducted the statistical modeling and geospatial risk mapping in Python and R, and drafted the original manuscript. Idorenyin Bassey Ekerette contributed to the study design, assisted in the validation and synthesis of the entomological data indices, and critically reviewed and edited the manuscript for intellectual content. Both authors read and approved the final version of the manuscript.

## Acknowledgements

The authors thank the many Nigerians and international entomologists whose published field surveys made this synthesis possible. We also thank the Natural Earth project for providing public-domain administrative boundary data.

